# Well, or Well Enough? An exploration of the need for guidance addressing clinician wellbeing and fitness to practice in UK Clinical Psychologists

**DOI:** 10.1101/2025.02.03.25321570

**Authors:** L. Birkett, A. J. Hagan, S.J. Verity

## Abstract

Practitioner Psychologists in the UK are required to meet standards of practice pertinent to their professional grouping to maintain registration with the Health and Care Professional Council. Updated guidance in effect from 1^st^ September 2023 states that registrants are now required to develop and adopt clear strategies for physical and mental self-care, thus requiring registrants to be agentic in implementing active methods of well-being management. A survey of 301 UK based Clinical Psychologists sought to determine whether a self-assessment tool to identify risk factors for wellbeing would be of benefit. The majority of participants endorsed the potential utility, necessity, and benefit of such a measure. Limitations of a self-assessment wellbeing measure are discussed, as well as systemic issues that impact well-being. This paper supports the development of a self-assessment wellbeing tool for those working within clinical psychology.

## Introduction

The term ‘well-being’ encompasses all aspects of physical and mental wellness. The APA Dictionary of Psychology defines this as ‘a state of happiness and contentment, with low levels of distress, overall good physical and mental health and outlook, or good quality of life’ (American Psychological Association, 2015). The maintenance of well-being is a condition of continued professional registration for UK-based practitioner psychologists. The Health and Care Professions Council (HCPC) Standards of Proficiency for Practitioner Psychologists state that registrants must: “3.2 [U]nderstand the importance of maintaining their own health” and “3.4 [B]e able to manage the physical, psychological and emotional impact of their practice” (Health & Care Professions Council, 2015). Updated guidance in effect from 1^st^ September 2023 states that: “[R]egistrants are now required to develop and adopt clear strategies for physical and mental self-care” (Health & Care Professions Council, 2022). Changes in phrasing demonstrate progress towards the requirement for active engagement with standards rather than solely passive assent, and that methods employed in maintenance of well-being should be well-defined, and transparent. The movement towards active management of well-being corresponds with the British Psychological Society (BPS) Code of Ethics that states the need for members to consider “[…] limitations to their competence *taking mitigating actions as necessary*” (British Psychological Society, 2021) (authors own emphasis).

Maintenance of personal well-being can present a challenge to clinical psychologists. Although compassion and empathy for patients is taught as a pillar of psychological care. Mental health workers are shown to have less self-compassion, leading to poorer well-being (Jena, Pradhan & Panigrahy, 2018). Practitioners are subject to multiple well-documented occupational risk factors that can impair well-being, including emotional ‘burnout’, compassion-related fatigue, and secondary trauma (Canfield, 2005; Figley, 2002; McCormack et al., 2018). In a study of mental well-being, 29.6% of clinical and counselling psychologists reported moderate levels of emotional exhaustion (Simpson et al., 2018). The poor well-being is often noted in the literature in the form of burnout as this is arguably the most notable and tangible output of poor mental health.

In addition to the existing occupational challenges to psychologists’ well-being, practitioners encounter the consequences of the COVID-19 pandemic. The continued phenomenon of the pandemic has a measurable impact on the well-being of many individuals with a greater negative impact on those experiencing pre-existing mental health inequalities (Di Gessa et al., 2021). Psychologists are not, of course, exempt from the distress inherent in the pandemic experience. Indeed, over 56% of psychologists responding to the study of Cerasa et al. (2022) reported the pandemic to have negatively affected their mental health. Practitioners were obliged to make immediate adjustments to support continued service provision for a disproportionately vulnerable patient population. Rather uniquely, healthcare professionals found themselves in the position of advising on and providing pandemic health support whilst simultaneously immersed in the pandemic experience. Despite the generationally unprecedented situation, psychologists made a substantial early contribution to the pandemic literature offering psychologically informed insight into the needs of the population (see O’Connor et al., 2020 amongst others). Coming into the fourth year of the pandemic, both adult and child mental health services record an unprecedented trajectory of increased referrals since pre-pandemic (Department of Health and Social Care, 2023). Referrals for emergency child mental health trebled between May 2019 and May 2023 (NHS Digital, 2023). The need for adult mental health services have also grown over this period, albeit at a slightly slower rate, with mental health service referrals showing a 22% increase per year compared to pre-pandemic (NHS Digital, 2023). These data suggest that psychologists, in common with other mental healthcare professionals, face increased pressure to meet the need of our rapidly growing patient population.

Given the occupational and systemic factors that risk the well-being of psychologists, it is important to identify methods of support for members of the profession. As the maintenance of well-being forms a condition of professional registration and mitigation of risk to clinical competence becomes individual responsibility, a self-assessment tool with the potential to identify risk to well-being may serve a valuable purpose. A number of tools already in existence may meet this need. Hirst & Nash researched the prevalence of burnout in social work, creating the Professional Wellbeing Self-assessment Tool (Hirst, 2019). The Professional Wellbeing Self-assessment Tool is a reflective tool that addresses seven dimensions of well-being, these being derived from a literature review into the key risks to well-being in social work. The NHS Leadership Academy offer a new online self-assessment tool for well-being for health and social care workers over the age of 18 years (NHS England, 2023). This tool provides a simplified version of a psychological battery of self-report questionnaires including the GAD-7 and PHQ-9 to offer the user areas they may benefit from intervention in.

This study presents data on the experience of clinical psychologists negotiating well-being in the context of their professional setting. Our primary aim was to explore the potential utility of a profession-specific assessment tool to support psychologists to make fitness to practice decisions around well-being.

## Method

### Design

The current cross-sectional study used a purpose-developed questionnaire hosted on the online platform Survey Monkey between 15 January 2021 and 22 January 2022 (SurveyMonkey Audience, 1999).

### Participants

Practitioner Psychologists registered with the Health and Care Professions Council (HCPC) and based within the United Kingdom were invited to participate. 301 participants responded to the online questionnaire.

### Materials

A purpose-developed 9-point questionnaire was used to explore the potential interest and benefit of a clinician-led self-use tool to support clinicians making fitness to practice decisions for themselves (Appendix A). The questionnaire assessed three core areas; a participant’s ability to assess fitness to practice, their feelings regarding seeking support when they perceive fitness to practice may be impaired, and the perceived utility/relevance of a fitness to practice tool. Respondents were given the option not to answer any question.

### Procedure

Eligible participants were identified through an online recruitment strategy. Participants were sought by email contact with individual NHS Trusts via Heads of Department of Psychology, via the Facebook UK Clinical Psychology Group, and via email contact with Special Interest Groups. Once participants accessed the study link they were presented with an information sheet and consent statement. Those choosing to participate proceeded to the web-based questionnaire. On completion, data were automatically saved on the Survey Monkey platform. Contact details of one author (***) were provided on the Survey Monkey platform should participants wish to discuss the study further or wish to withdraw. No incentive was given to individuals to participate. Demographic data on participants’ stage of career and context of practice were not gathered at initial data collection.

### Analysis

Quantitative data analysis is used to provide descriptive outcomes of questions one to seven. Qualitative analysis utilised the grounded theory approach (Charmaz, 2006). Grounded theory analysis is based on an inductive approach that seeks to explore meanings made and narratives constructed within qualitative data. Following familiarisation with the data, themes were identified and labelled as per Braun & Clark (2006). The coding system was ratified by a second co-rater to ensure accuracy, relevancy, and reduce bias (O’Connor & Joffe, 2020). Data saturation was established by ensuring all data could be coded to one of the themes identified and no further fields felt necessary to encompass the information conveyed in the data. Final analyses were shared with a sample of participants as a member check. The anonymised data are available from the corresponding author upon reasonable request.

### Ethical Considerations

The survey was sponsored by Newcastle University (Reg. PIF 4019) with Newcastle Upon Tyne Hospitals NHS Foundation Trust registered as the host site for the research (Registration No. 13755). Participants gave informed consent for their data to be anonymised and collected by completion of the survey as outlined in the participant information sheet (see Appendix A). Information for participants distressed by their participation was provided in the body of the survey.

## Results

A total of 301 Health and Care Professions Council (HCPC) registered UK psychologists participated in the current study.

### Assessing fitness to practice

Of the 299 responses to question one (‘Have you received any formal training in how to assess fitness to practice’), 258 clinicians (86%) reported having received *no training* on how to assess their fitness to practice with respect to wellbeing (see Figure 1).

**Figure 1.**
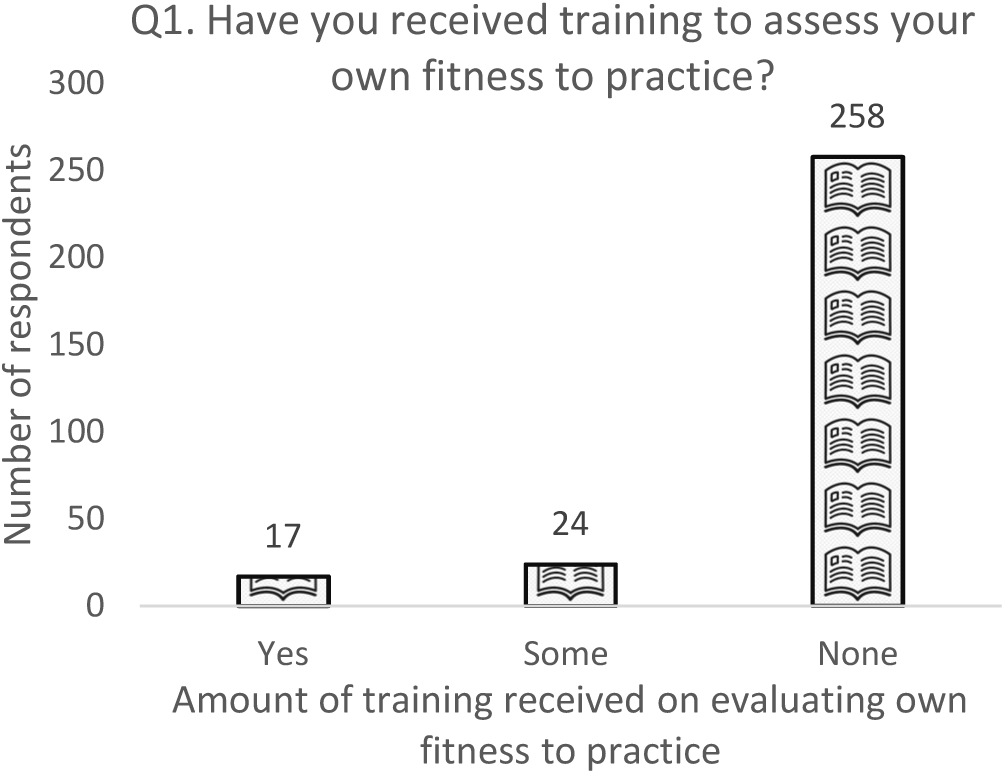
Number of clinicians who have received training on assessment of their own fitness to practice.

Question two asked participants to rate their level of confidence when assessing their own fitness to practice, or that or a supervisee. On a scale of 0-10 (0 = no confidence, 10 = fully confident), participants rated their confidence at an average of 5.5.

### Seeking appropriate support

Questions three and four assessed the likelihood of speaking to a supervisor about their current poor mental wellbeing, and their level of comfort/discomfort in doing so. In terms of likelihood of speaking to a supervisor, 176 participants (58.6%) reported that they *would speak* to their supervisor (Q3). In comparison 90 participants (30%) reported that they would be *very unlikely* or *unlikely* to speak with their supervisor if experiencing poor wellbeing (Figure 2).

**Figure 2.**
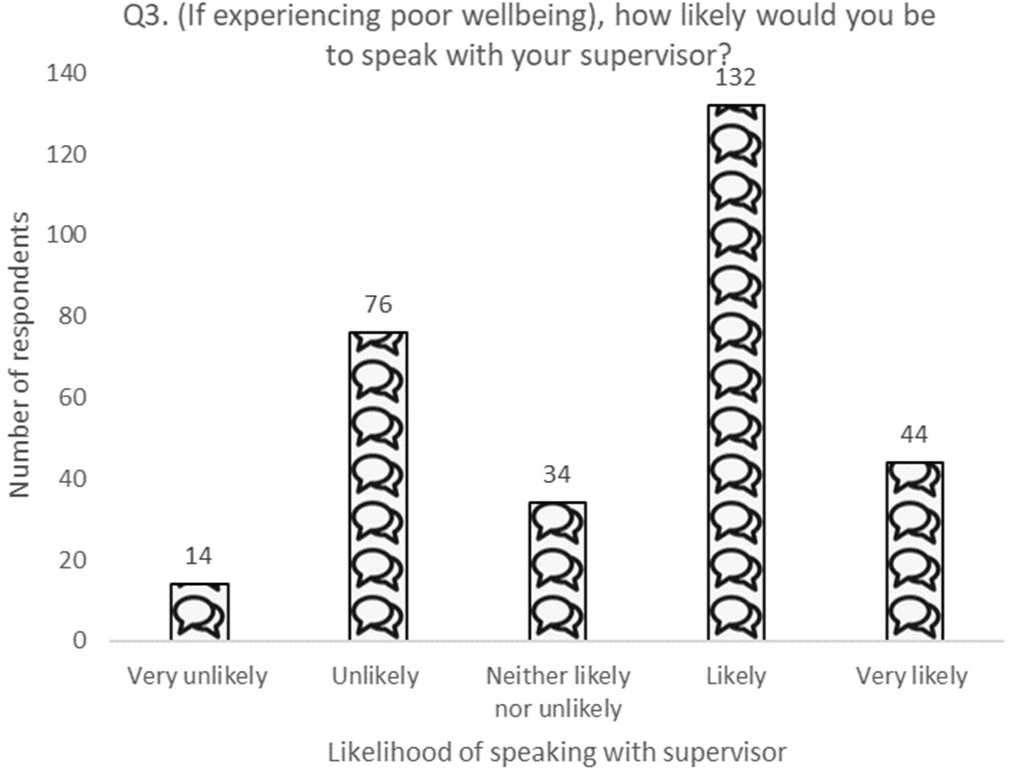
The likelihood of a clinician speaking to their supervisor regarding their well-being.

In terms of comfort/discomfort, 68 participants (22%) reported that they would feel *very comfortable* or *extremely comfortable* in speaking to a supervisor regarding their well-being (Q4). In comparison 141 participants (46.8%) stated they would feel *rather* or *extremely uncomfortable* (Figure 3).

**Figure 3.**
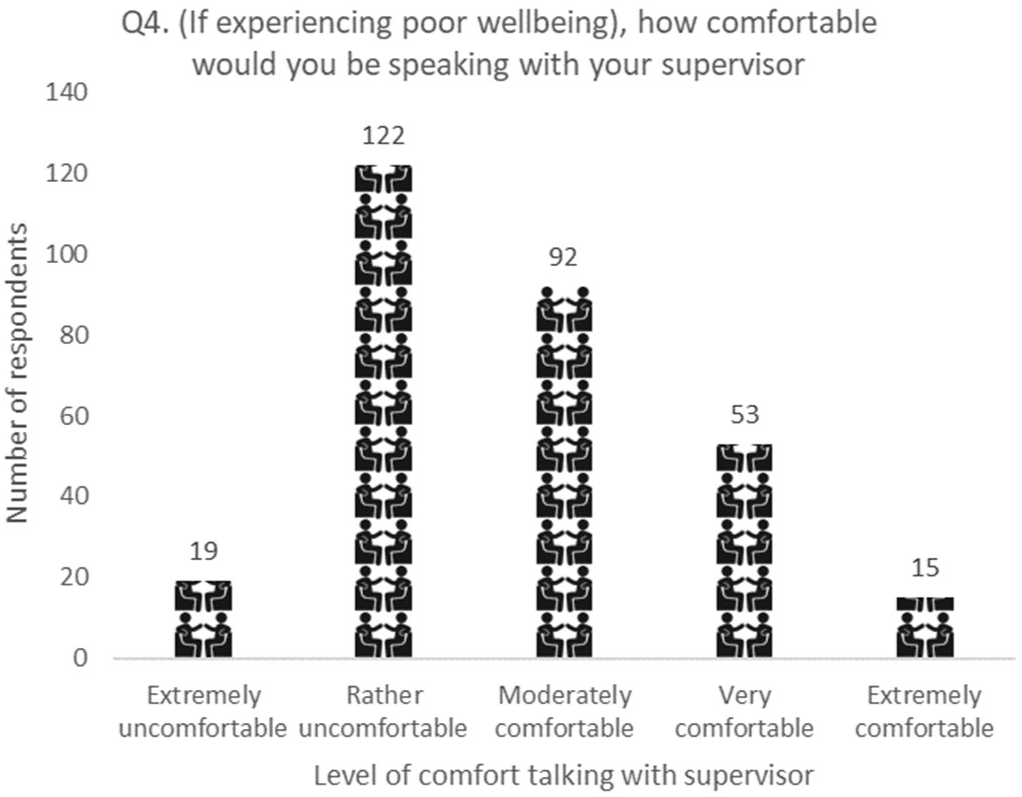
How comfortable a clinician would feel speaking to their supervisor regarding their well-being.

### The usefulness & relevance of a fitness to practice measure

Questions five and six assessed the perceived usefulness (Q5) and relevance (Q6) of a self-assessment fitness to practice tool. Of the 299 responses to question 5, 213 (71%) respondents felt such a tool would be *very* or *extremely* useful (Figure 4).

**Figure 4.**
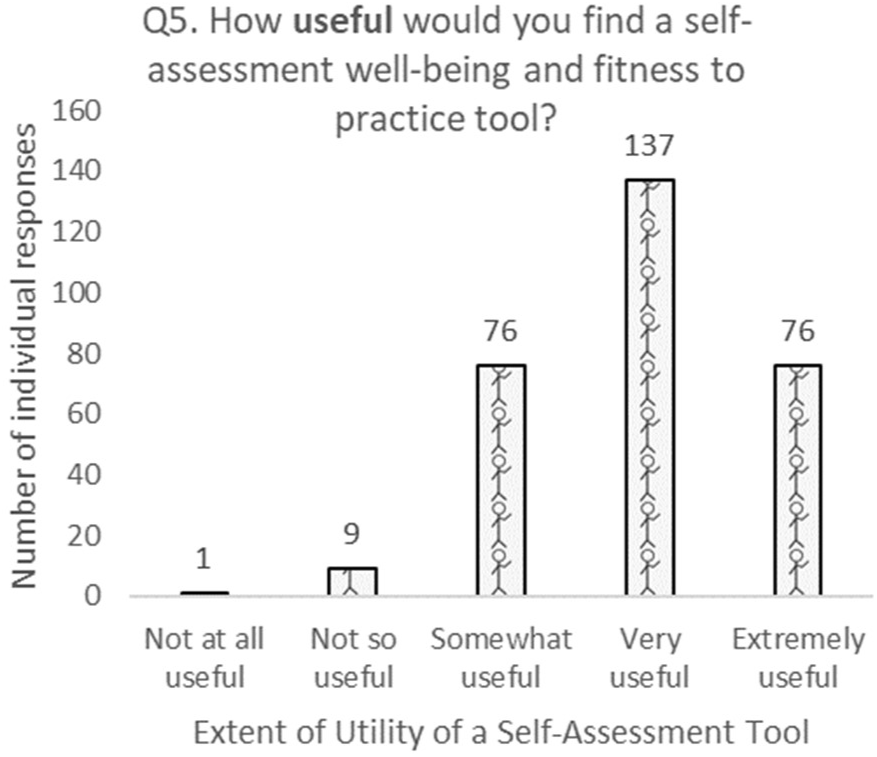
How useful a self-assessment tool may be.

In terms of relevance, 203 (67%) of 301 respondents felt that a self-assessment tool might be *very* or *extremely* relevant to them (Figure 5).

**Figure 5.**
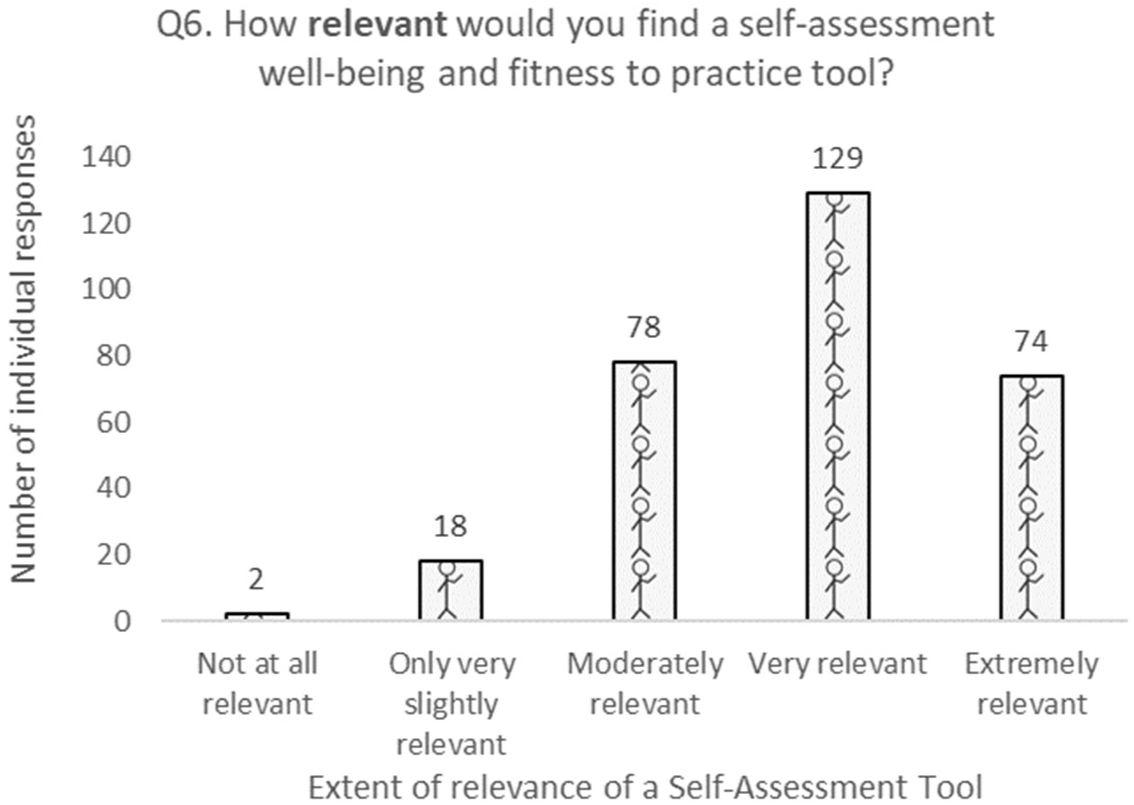
How relevant a self-assessment tool may be.

### Lived experiences

Responses to question seven (‘personal experience with mental health condition, stress or distress’) indicate 164 participants (79%) had experienced a mental health condition, stress, or distress (Figure 6). Only 5 (3%) of these participants felt sure about whether to take time off and/or seek help during this period.

**Figure 6.**
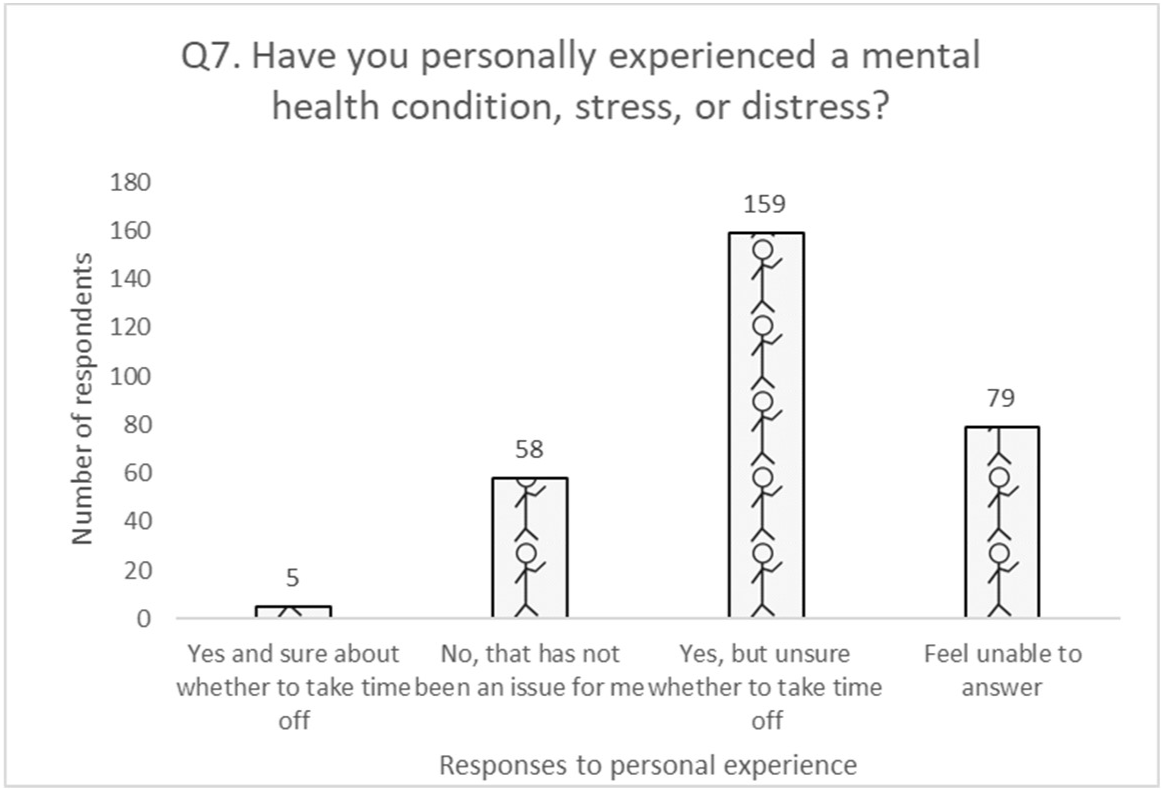
Lived experience of either a mental health condition, stress, or distress

### Qualitative data

Question nine provided participants with a free text space to provide any additional comments, of which 65 participants (21.6%) provided a response. Initial coding of the data suggested five overarching themes: Fitness to practice, Current Support, Stigma, Barriers for Support, and Limitations of Measure.

**Figure 7.**
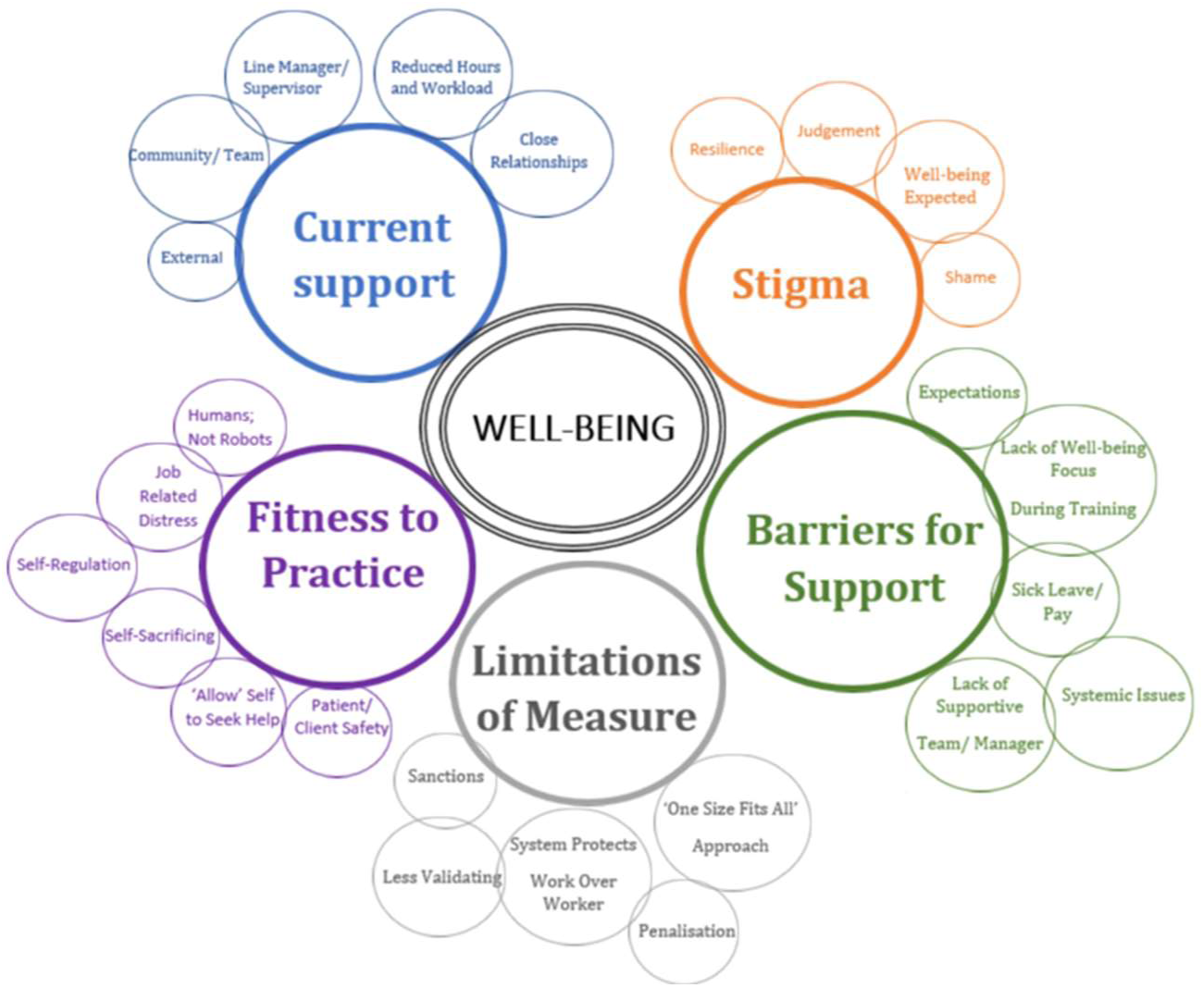
Illustration of hierarchical themes

### Fitness to practice

The theme of ‘fitness to practice’ encompasses responses associated by participants with the notion of being ‘fit to practice’. This theme included sub-themes such as: ‘being self-sacrificing’, ‘experiencing job-related distress’, ‘allowing oneself permission to’ seek help’, ‘[keeping well to support] patient safety’, and ‘self-regulation’. Examples of data placed in this category are: *“…FTP is both about … welfare and responsibility to clients”*.

### Limitations of measure

This theme discusses the potential drawbacks and limitations regarding the development and implementation of a measure aimed at assessing fitness to practice. Identified sub-themes were: ‘potential sanctions’, ‘[feeling that the] system protects work over worker’, ‘[fear of] penalisation for not being well’, and the perception of a ‘one-size-fits-all’ institutional approach to wellbeing and fitness to practice. Examples of data placed in this category are: *“[I wonder]… how might it be used and interpreted i.e. by HR”, and “Uncertainty about how disclosing this could impact on their registration with regulatory bodies”*.

### Barriers to support

The theme of ‘barriers to support’ addresses the reasons individuals do not seek support. Sub-themes identified were: ‘systemic issues’, ‘lack of wellbeing focus during training’, ‘sick leave/pay’, ‘[perceived] expectations’, and ‘lack of a supportive team/manager’. Examples of data placed in this category were: *“[S]ome people might not have as supportive a management structure and in those cases it would be more difficult to feel comfortable sharing distress with them”*.

### Stigma

The theme of ‘Stigma’ refers to the stigma felt from the individual or others regarding accommodations to promote wellbeing. Sub-themes identified were: ‘[perceived] judgement’, ‘shame’, and ‘[perceived expectations of] resilience’. Examples of data placed in this category were*: “I’ve noticed an assumption that psychologists will be ready, able, and have limitless capacity to support everyone”*; *“[O]ur own support and wellbeing was somewhat assumed”*; and *“[…] still a long way to go with people feeling free to speak out”*.

### Accessible support

The theme of ‘Accessible support’ addresses the perceived accessibility of current support networks. Sub-themes identified were: ‘external support’, ‘community/team support’, ‘line manager/supervisor relationship’, ‘reduced hours or workload’, and ‘relationships’. Data in this category included: *“[L]uckily for me I had/have excellent line management and supervision”,* and *“What helps is having someone safe you can trust and call”*.

## Discussion

The current study explores the mental health of clinical psychologists and polls the perceived utility of a self-use assessment tool to support making fitness to practice decisions. 301 Health and Care Professions Council (HCPC) registered clinical psychologists participated.

### Mental health experience of clinical psychologists

This study found 79% of participants to have personal experience of stress or distress that had negatively affected their mental health. This figure is perhaps unsurprising given the occupational pressures and systemic factors discussed earlier, personal factors notwithstanding. Only 3% of participants had felt sure about whether they should take time off work during a period of poor mental health, stress, or emotional distress, whilst 86% reported uncertainty about whether to take time off. When experiencing poor mental health, 30% of participants reported that they would be very unlikely or unlikely to talk to their supervisor about their mental wellbeing. It was raised during review of this manuscript that our question pertained only to ‘how likely’ the supervisee would be to talk to their supervisor about their mental health rather than asking more widely about supervisory or management support. Indeed, it is possible that the supervisees in question have a plethora of alternative routes of support offered within their professional contexts. Nonetheless, we would suggest that in meeting the demands of professional registration that the majority of registrants would consider it their duty to raise significant impact on mental health with their supervisor. The fact that so many participants would not be likely to do so requires at least some consideration of the systemic reasons discouraging these conversations.

### Potential reasons for reticence to use supervision to acknowledge and support mental health needs

Our data suggests that, even whilst able to identify the impact of occupational or personal factors upon mental health, psychologists do not make optimal use of the provision of supervision. The themes of stigma, shame, and perceived negative judgement towards clinicians experiencing mental distress were frequently identified via analysis of the questionnaire data. These findings are unsurprising. The American Psychological Association found that when experiencing thoughts of suicide, only 50% of clinicians were willing to talk about their difficulties. Garelick et al. (2012) identified that mental health professionals often feel a high degree of shame about their experience of poor health and are fearful of professional judgement. Closer to home, a study of clinical psychologists conducted via the British Psychological Society Division of Clinical Psychology found 62.7% of participants to report lived experience of a mental health difficulty (Tay et al., 2018). This figure is rather less than our own finding of 79% of respondents, but shows that a substantial proportion of respondents to online survey report difficulty. The same study found participants to speak of the perceived stigma of having a mental health condition, again, consistent with the findings of the current study. Despite the professional prioritisation of formulation over diagnostic labelling, it appears that clinical psychologists are not inoculated against damaging societal narratives about poor mental health.

### Perceived utility of self-assessment guidance or measures

The fact that there is no tool supporting the self-assessment of clinician well-being, yet there are clear institutional and regulatory repercussions for those who fail to do so effectively, suggests scope for improvement. Responses to the current study indicate a largely positive response to any potential guidance and/or a measure to assess well-being in regard to fitness to practice. 71% of participants felt that guidance and/or a measure would be very or extremely useful, and 67% felt that it would be very or extremely personally relevant.

### Limitations of a self-use measure of mental wellbeing and fitness to practice

Although the findings of this study are largely in support of a well-being measure, the potential limitations of such a measure must also be considered. The potential for a measure being used to further stigmatise and disempower clinicians was of concern to participants. A further limitation may relate to the message inherent in providing a ‘self-help’ tool. Whilst the autonomy offered by self-help measures offers benefit to some individuals, it may be perceived by others as a further effort by institutions and systems to leave individuals solely responsible for managing poor mental health (Simionato & Simpson, 2018). This is, perhaps, the most significant limitation. A self-help rating tool only allows clinicians to rate or describe their distress, not to change the systems around them that are the determinants of mental health.

The current level of psychologists employed in the UK is insufficient to meet the growing needs of the population (Branley & Byrne, 2012). Insufficient levels of professionals in the mental health arena mean that psychologists frequently work in understaffed hospitals or centres with untenable workloads of, increasingly, more complex patients who have themselves endured substantial wait times to receive an appointment. It is beyond the scope of the current study to address the political and social context of the current mental health provision in the UK. We must, however, acknowledge the need to find ways to safeguard the mental health of clinicians without placing responsibility on individuals to ‘survive’ an unsustainable system. Any tool, measure, or guideline is not proposed as an all-encompassing solution to a complex problem and should not be seen as a replacement for the necessary improvement of systemic issues.

### Strengths and limitations of the study

This study is subject to a number of methodological limitations. First, despite certain demographic groups being at higher risk of poor wellbeing (Hodes & Epperson, 2019; Vahdaninia et al., 2020), no participant demographic information was collected in this study. This obscures the known additional burden on mental wellbeing experienced by clinicians from, for example, Black and other ethnically minoritised groups. Further, it does not allow for difference in sex, area of work, or level of workload to be taken into account. The lack of demographic data also raises a threat to generalisation of these findings as the 301 participants may not be representative of the approximately 13,000 clinical psychologists on the HCPC register. Second, the use of a structured questionnaire with predetermined questions may have limited data gathered on this topic. Use of a fully exploratory methodology, such as semi-structured interview and use of qualitative analytic methods would add significantly to the evidence base. With this in mind future research design should foreground the knowledge and experience of clinicians who are representative of the wider population.

## Conclusion

The mental well-being of clinical psychologists is of individual, service-level, institutional, and societal importance. In a political climate of sustained healthcare underfunding, clinical psychologists are called upon to manage increasing numbers of patients with greater complexity. Despite regulatory requirements for clinical psychologists to take individual responsibility for the maintenance of their own fitness to practice (including mental health), there are currently no tools to support clinical psychologists to determine when mental health might be impacting upon fitness to practice. The current study demonstrates both a need for and positive appetite for development of a tool or guidelines to support clinicians experiencing poor mental health, stress, or distress.

## Data availability statement

The data that support the findings of this study are available from the corresponding author upon reasonable request.

## Acknowledgements

The author(s) received no financial support for the research, authorship, and/or publication of this article.

